# Oxytocin reduces subjective fear in naturalistic social contexts via enhancing top-down middle cingulate amygdala regulation and brain-wide fear representations

**DOI:** 10.1101/2024.04.22.24306196

**Authors:** Kun Fu, Shuyue Xu, Zheng Zhang, Dan Liu, Ting Xu, Yuan Zhang, Feng Zhou, Xiaodong Zhang, Chunmei Lan, Junjie Wang, Lan Wang, Jingxian He, Keith M Kendrick, Bharat Biswal, Dezhong Yao, Zhen Liang, Weihua Zhao, Benjamin Becker

**Affiliations:** The Center of Psychosomatic Medicine, Sichuan Provincial Center for Mental Health, Sichuan Provincial People’s Hospital, University of Electronic Science and Technology of China, Chengdu, China; MOE Key Laboratory for Neuroinformation, School of Life Science and Technology, University of Electronic Science and Technology of China, Chengdu, China; School of Biomedical Engineering, Medical School, Shenzhen University, Shenzhen, China; Guangdong Provincial Key Laboratory of Biomedical Measurements and Ultrasound Imaging, Shenzhen, China; Faculty of Psychology, Southwest University, Chongqing, China; Key Laboratory of Cognition and Personality, Ministry of Education, Chongqing, China; Department of Biomedical Engineering, New Jersey Institute of Technology, Newark, USA; State Key Laboratory of Brain and Cognitive Sciences, The University of Hong Kong, Hong Kong, China; Department of Psychology, The University of Hong Kong, Hong Kong, China

**Keywords:** Oxytocin, Fear, Anxiety, fMRI, Amygdala

## Abstract

Accumulating evidence from animal and human studies suggests a fear-regulating potential of the neuropeptide oxytocin (OT), yet the clinical translation into novel interventions for pathological fear requires a behavioral and neurofunctional characterization under close-to-real life conditions. Here, we combined a naturalistic fMRI-design inducing high and immersive fear experience in social and non-social contexts with a preregistered between-subjects randomized double-blind placebo-controlled intranasal OT trial (24 IU, n = 67 healthy men). OT selectively reduced subjective fear in social contexts but not in non-social contexts. Specifically, OT enhanced left middle cingulate cortex (lMCC) activation and its functional connectivity with the contralateral amygdala, with both neural indices significantly and inversely associated with subjective fear following OT. On the network level, OT enhanced communication between the dorsal attention network (DAN) with the fronto-parietal (FPN) and the default-mode network (DMN) as well as on the more fine-grained level brain-wide communication. Utilizing an independent activity-connectivity neuromarker for fear in naturalistic contexts (CAFE) confirmed that OT attenuated brain-wide fear expressions. Findings indicate an ecologically valid and social-specific fear-reducing potential of OT, highlighting its potential value as a treatment option for disorders characterized by excessive fear in social situations.

## Introduction

Fear is an evolutionarily conserved and highly adaptive defensive avoidance response, fundamental to surviving and functioning in everyday life ^1–4^. The experience of excessive and overwhelming fear represents a hallmark symptom of anxiety and trauma-related disorders ^5–7^. These highly prevalent psychiatric disorders have become a leading cause of disability and are accompanied by personal suffering as well as an enormous socio-economic burden. The conventional pharmacological interventions targeting the classical neurotransmitter (GABA-ergic, serotonergic) systems 7-9 are characterized by moderate response rates and potential negative side effects ranging from dizziness towards addiction. While several promising candidate compounds have shown effects in animal models, no new pharmacological treatment for fear or anxiety has entered the clinical practice during the last decades (for recent non-pharmacological developments see e.g. ^10,11^).

Based on promising findings in animal models and preclinical human studies neuropeptides, including oxytocin (OT), vasopressin or angiotensin II, have been suggested as promising new strategies to regulate fear and anxiety ^12–16^. Most previous research in this field has focused on OT, a neuropeptide produced in the hypothalamus, and animal models 15,16 as well as preclinical human proof-of-concept studies demonstrated promising effects of OT on regulating negative emotional processing in the domains of fear and anxiety 16-20. Some of the effects of OT on cognitive and affective domains appear to be specific or at least pronounced in the social context. Accumulating evidence from animal models suggests that OT regulates the detection and evaluation of social cues via neural circuits involved in social salience processing^21–23^. In humans, overarching frameworks suggest that oxytocin enhances the salience of social information, exerting a person- and context-dependent modulation of responses towards both positive and negative social cues^24–26^. In support of social context-dependent effects of OT, previous human studies reported that exogenous OT decreases top-down attention regulation towards social but not non-social stimuli^27,28^ and selectively modulates learning from and approach towards social stimuli^29–32^. However, other studies also reported common behavioral effects of OT on social and non-social stimuli for instance in reward- or avoidance-related domains^26,33^.

OT neurons from the hypothalamus project to a wide range of brain regions involved in fear, anxiety as well as social processes, including the amygdala, hippocampus, midbrain and frontal lobe^5,34^, and previous studies indicate that intranasal OT can regulate these processes via influencing activity in the amygdala, dorsal anterior cingulate cortex (dACC) and medial prefrontal cortex7,18-20,35-37. Previous studies have demonstrated that intranasal OT can attenuate subjective anxiety and enhance regulation over negative emotions in individuals with high trait anxiety^38,39^. On the neural level, OT attenuated amygdala reactivity towards threat-related stimuli7,18,40 and modulated insula activity in response to negative and threat-associated stimuli^41,42^ as well as the interaction between these regions43. Cortical midline regions, including cingulate regions and medial frontal regions, exhibited modulation via OT during a number or fear-associated paradigms, including threat conditioning^44,45^. Initial clinical studies in patient populations moreover demonstrated that OT normalizes amygdala-insula and amygdala-mid cingulate cortex (MCC) communication in generalized social anxiety disorder^36^, suggesting that OT may not only regulate activity in isolated regions but may additionally exert its effects via modulating functional connectivity between these regions. However, despite the initially promising findings from these studies, the fear and anxiety-reducing effects of OT and the underlying neural circuits have been determined under laboratory experimental contexts that strongly differ from the dynamic contexts during which fear arises in everyday life ^46^. Specifically, these studies combined the intranasal administration of OT with sparsely presented static experimental stimuli (e.g. pictures of fearful faces or scenes) and primarily focused on treatment-induced changes in brain regional activity during fMRI.

This approach stands in contrast to contemporary neuroscience models proposing that fear emerges in interaction between subjective appraisal and dynamic changes in the environment ^47,48^ and that the corresponding mental states are mediated by an intricate interplay among distributed yet interacting brain systems 49-51 To account for the complexity of subjective affective experiences under ecologically valid conditions and to capture the underlying network-level communication in the brain, recent overarching conceptualizations have proposed the combination of movie stimuli as highly immersive and dynamic stimuli with network-level analyses 52,53. Initial empirical studies confirmed that these stimuli can induce highly immersive affective experiences in complex social situations ^54,55^ as well as the essential role that network-level communication plays in establishing affective and social experiences under naturalistic contexts ^46,53,56,57^.

Mental disorders, including fear-related disorders, have been reconceptualized as network-level disorders that are characterized by dysregulated regional activity as well as network level connectivity (putatively reflecting communication between brain regions) ^51,58,59^. Together with a growing number of studies underscoring the potential of OT to regulate neurofunctional network-level interactions in fear-related networks across species ^60–63^, which may be partly explained by the physiological properties of the oxytocin-signaling system, including an axonal release, widespread receptor distribution and considerably longer half-life compared to classical neurotransmitters ^64^, these findings indicate the necessity to account for network-level effects to fully determine a regulatory effect of OT on fear experience and its therapeutic potential.

Against this background, the present pre-registered between-subjects randomized double-blind placebo-controlled pharmaco-fMRI study (n = 67 healthy male participants) capitalized on recent progress in naturalistic fMRI and network-level analyses to determine whether a single dosage of intranasal OT (1) reduces the subjective experience of fear under highly immersive and ecologically valid conditions, (2) modulates concomitant regional activity in brain systems identified in previous studies, (3) modulates brain communication of the identified regions and on the whole-brain network level, and (4) effects vary as a function of social context. Based on previous studies 7,18-20, we primarily hypothesized that intranasal OT would decrease subjective fear in naturalistic contexts with more pronounced effects in the social domain while concomitantly enhancing neural activity in regulatory brain regions and inhibiting activity in fear-processing brain regions. To further characterize these effects, exploratory analyses examined changes in functional connectivity, brain-wide communication patterns, and responses of a recently developed multivariate neuromarker for fear in dynamic naturalistic contexts (CAFE^46^). Based on prior research, we hypothesized that OT would modulate connectivity, including amygdala connectivity, influence large-scale brain network interactions, and attenuate the expression of the CAFE fear signature^46,65,66^.

## Results

### Demographics and potential confounders

The OT (n = 33) and PLC (n = 34) groups were comparable with respect to socio-demographics, mood and mental health indices arguing against nonspecific treatment effects (**Table 1**, all *ps >* 0.07).

**Table 1.**
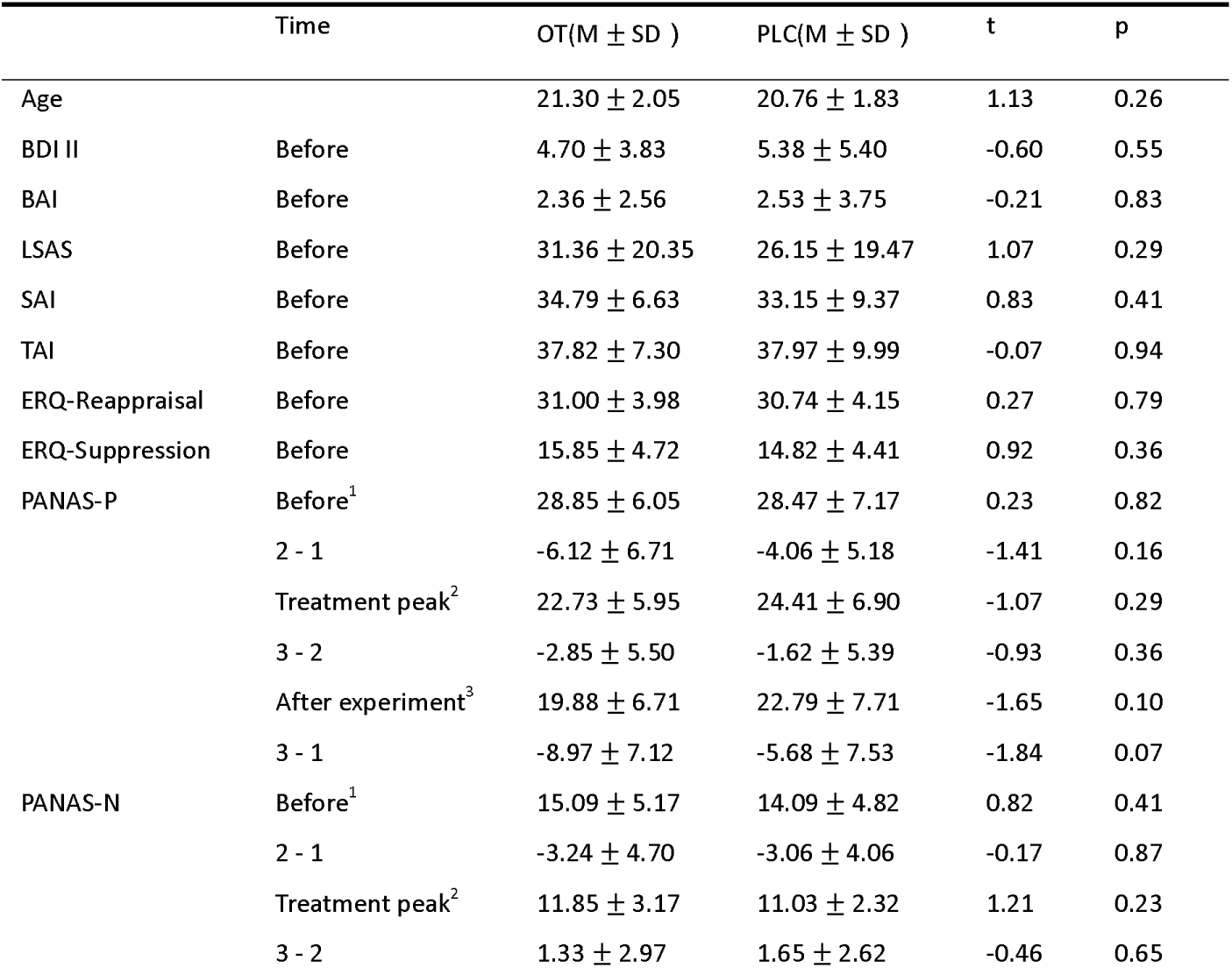
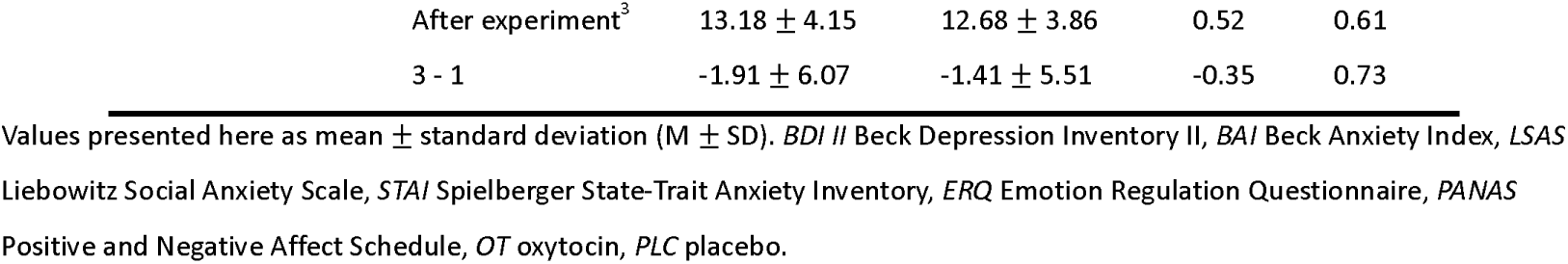
Demographics and potential confounders in the two treatment groups.

### OT reduced subjective fear experience in social fear naturalistic contexts

The ANOVA revealed a significant main effect of emotion (*F*_(1,65)_ = 1434, *p* < 0.0001,η_p_^2^ = 0.957, **Fig. 1a**) such that participants rated subjective fear considerably higher during fear trials (FS and FNS) than neutral ones, and a significant main effect of social (*F*_(1,65)_ = 76.81, *p* < 0.0001,η_p_^2^ = 0.542, **Fig. 1a**) reflecting lower fear during social trials (FS and NS) compared with non-social trials, and a marginally significant main effect of treatment (*F*_(1,65)_ = 3.31, *p* = 0.074, η_p_^2^ = 0.048, **Fig. 1a**). The effects of OT were further claified by a significant interaction effect between treatment and emotion (*F*_(1,65)_ = 4.064, *p* = 0.048, η_p_^2^ = 0.059, **Fig. 1a**), while the three-way interaction effect (treatment × emotion × social) failed to reach significance (*F*_(1,65)_ = 0.869, *p* = 0.355, **Fig. 1a**). To further determine the effects of OT we conducted exploratory post-hoc tests in the different social contexts, and the results indicated that OT significantly decreased fear experience in response to fearful stimuli in naturalistic social contexts but not in non-social contexts (*t*_(1,65)_ = -2.252, *p* = 0.028, Cohen’s *d* = -0.55 under FS while *t*_(1,65)_ = -1.475, *p* = 0.145, Cohen’s *d* = -0.36 under FNS, **Fig. 1b**). Finally, the interaction between emotion and social was significant (*F*_(1,65)_ = 53.15, *p* < 0.0001,η_p_^2^ = 0.45, **Fig. 1a**) reflecting that participants reported generally lower fear experience in social fear contexts. The main effect of treatment and other interactions were not significant. Together the results partly support that OT can reduce subjective fear in naturalistic social fear situation.

**Fig. 1.**
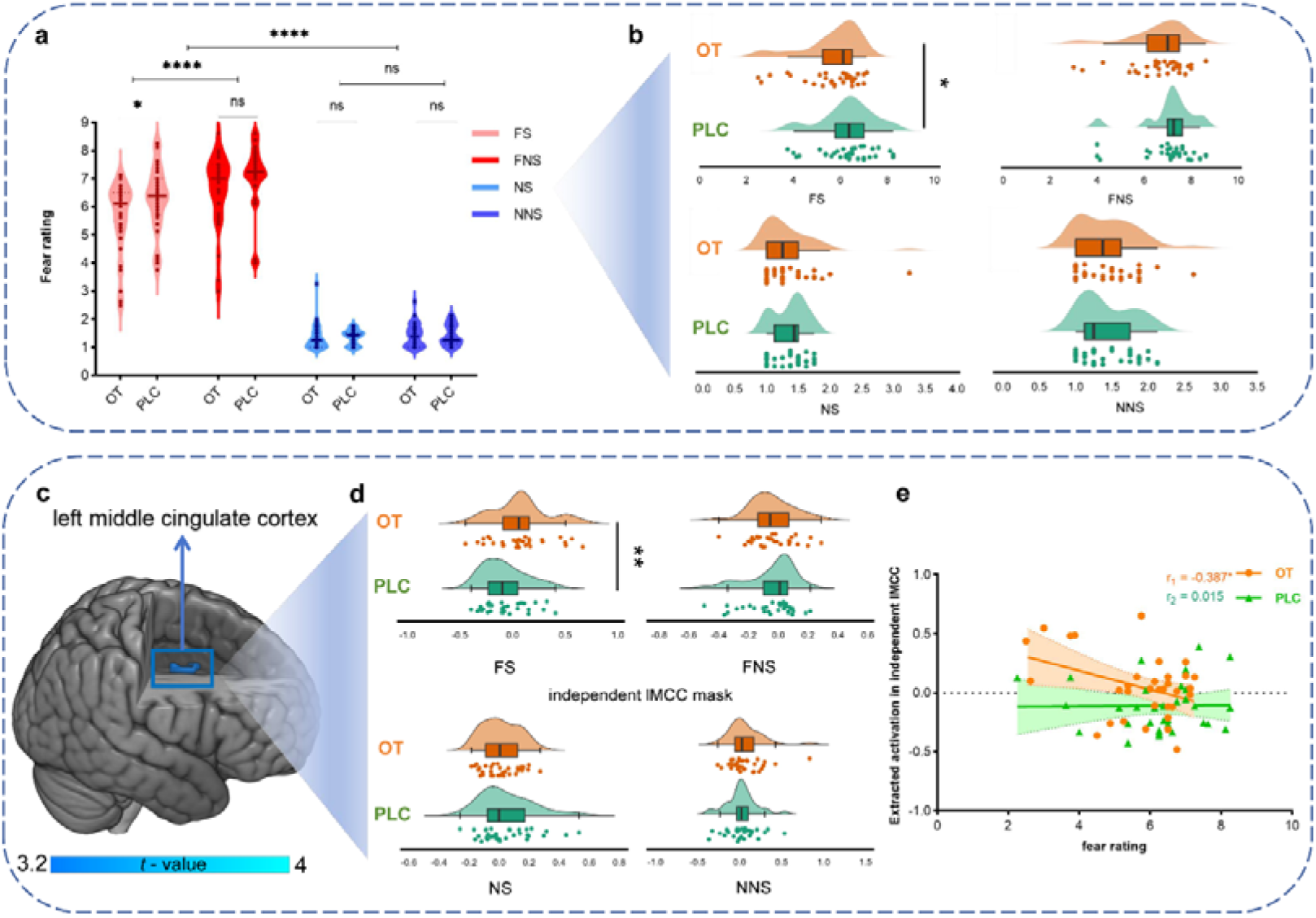
The effects of OT on behavioral fear rating and cingulate activity. **a** OT reduced subjective fear rating in social fear naturalistic situation, **b** OT effects on fear rating under four conditions separately, **c** OT but not PLC could strengthen the activation in the left middle cingulate cortex (lMCC) under the interaction of valence and social. Particularly, **d** OT specifically increased the activation in lMCC under FS but not other conditions. **e** The activation in lMCC was significantly negatively correlated with subjective fear experience in OT group but not PLC group. Please note that the group differences based on the extracted blood oxygenation level dependent (BOLD) signals in each of the separate conditions were presented for display purpose. OT oxytocin, PLC placebo, FS fear social, FNS fear non-social, NS neutral social, NNS neutral non-social. **** p_FDR-corrected_ < 0.0001, ** p_FDR-corrected_ < 0.01, * p_FDR-corrected_ < 0.05, ns not significant.

### OT enhanced middle cingulate activation in the social fear contexts

We initially examined brain regions participating in fear processing under naturalistic contexts with different social features compared with neutral control conditions, a series of corresponding voxel-wise one-sample t-tests conducted in the PLC group confirmed previous studies, suggesting that visual and temporal regions were strongly engaged during exposure to dynamic fear movie clips irrespective of social features (**Fig. S3**). Examining treatment effects revealed a significant interaction between treatmen × emotion × social located in the left middle cingulate cortex (peak MNI, x, y, z = 7, -3, 30, *t*_(1,65)_ = 67, K = 33, *p_FWE-peak(svc)_* =.019, **Fig. 1c**), with post-hoc t-tests on extracted activity measures from independently defined lMCC (FPN_Cing_1 from the Schaefer atlas^67^) indicating that compared to PLC, OT selectively enhanced activation in this region for FS (*t*_(1,65)_ = 2.69, *p* = 0.009, **Fig. 1d**) but not other conditions (all *p*s *>* 0.05, **Fig. 1d**). Examining associations between lMCC activity (extracted parameter estimates from the atlas region) and subjective fear experience revealed a significant negative association in the OT (*r* = -0.387, *p* = 0.026, **Fig. 1e**) but not in PLC group (*r* = 0.015, *p* = 0.934, **Fig. 1e**). A Fisher r-z transformation indicated that these correlation coefficients were significantly different between the OT and PLC groups (Z = -1.65, one-tailed *p* = 0.0495). It should be noted that the correlation analysis results between lMCC activation and subjective ratings need caution while interpret.

### OT enhanced MCC-amygdala functional communication consistently in the social fear contexts

Examining OT effects on the functional communication of the lMCC revealed a significant interaction effect between treatment *X* emotion *X* social located in the right amygdala (rAmy) (peak MNI, x, y, z = 22, -7, 13, *t*_(1,65)_ = 3.81, k = 19, *p_FWE-peak(svc)_=* 0.021, **Fig. 2a**), with post-hoc t-tests indicating that OT relative to PLC selectively enhanced lMCC-rAmy coupling in social fear contexts (*t*_(1,65)_ = 2.69, *p* = 0.009, **Fig. 2b**) but not other contexts (all *p*s *>* 0.05, FDR-corrected, **Fig. 2b**), confirming social fear-specific effects of OT on the neural level. Given that the specific associations between MCC-amygdala functional coupling and anxiety scores68, as well as the coupling was selectively modulated by OT rather than PLC^36^, we hypothesized that the associations between participants’ MCC-amygdala functional coupling and fear ratings would be different across treatment groups. We found that lMCC-rAmy functional coupling was significantly negatively associated with subjective fear experience in the OT (*r* = -0.426, *p* = 0.013, **Fig. 2c**) but not PLC group (*r* = -0.026, *p* = 0.883, **Fig. 2c**, Fisher *r*-to-*z* transformation revealed a significant difference between the treatment groups, Z = -1.68, one-tailed *p* = 0.0465).

**Fig. 2.**
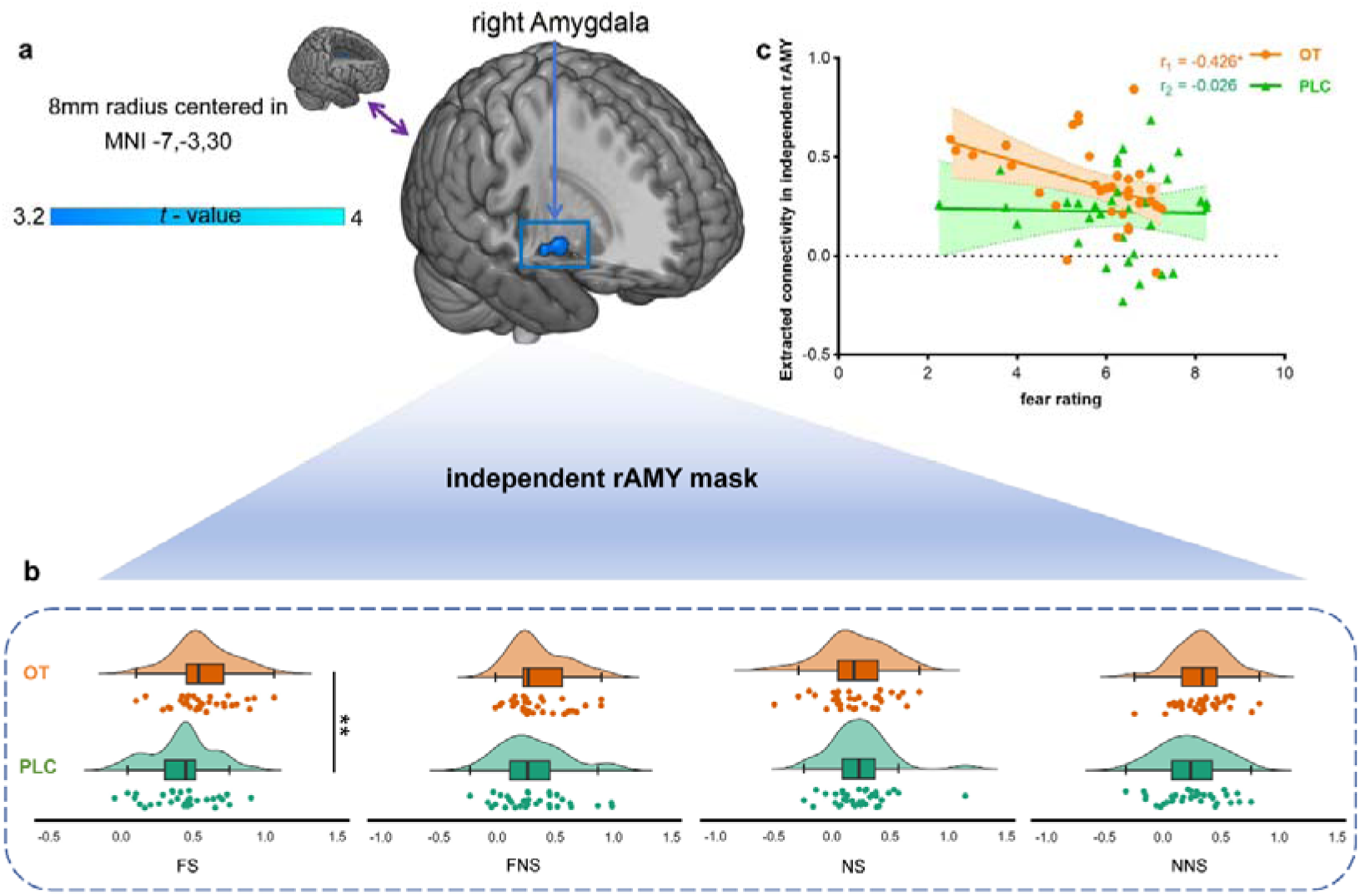
OT strengthened lMCC-rAmy communication specifically in social fear contexts. **a** Interaction effect between treatment X emotion X social on lMCC connectivity with the right amygdala **b** OT specifically increased functional connectivity between lMCC and rAmy in the FS contexts but not other conditions and c The functional connectivity between lMCC and rAmy was significantly negatively correlated with subjective fear experience in OT group but not PLC group. Please note that the group differences based on the extracted BOLD signals in each of the separate conditions were presented for display purpose. OT oxytocin, PLC placebo, FS fear social, FNS fear non-social, NS neutral social, NNS neutral non-social. ** pFDR-corrected < 0.01, * pFDR-corrected < 0.05, # pun-corrected < 0.05.

### OT enhanced FPN-DAN and DMN-DAN coupling during social fear contexts

Examining the effects of OT on the large-scale network level using separate two-sample t-tests on the matrices of z-values for the two main contrasts of interest (FS - NS, FNS - NNS) and following mantel test revealed that OT significantly affected network functional connectivity differently under two contexts (rho_mantel test_ = -0.1333, *p >* 0.05). Specifically, compared to PLC (all *p*s *>* 0.05, **Fig. 3f**), OT significantly enhanced the functional communication between FPN and DAN (*t*_(1,65)_ = 2.75, *p* = 0.0077, Cohen’s *d* = 0.672, **Fig. 3e**) as well as between DMN and DAN (*t*_(1,65)_ = 2.02, *p* = 0.048, Cohen’s *d* = 0.493, **Fig. 3e**) under the social fear but not the non-social fear contexts. The effects were confirmed by further analyses examining the interaction contrast ([FS – NS] – [FNS – NNS]), with results specifically underscoring that the network level effects of OT depend on the social versus non-social context (detailed methods and results please see Supplemental Methods and Results).

**Fig. 3.**
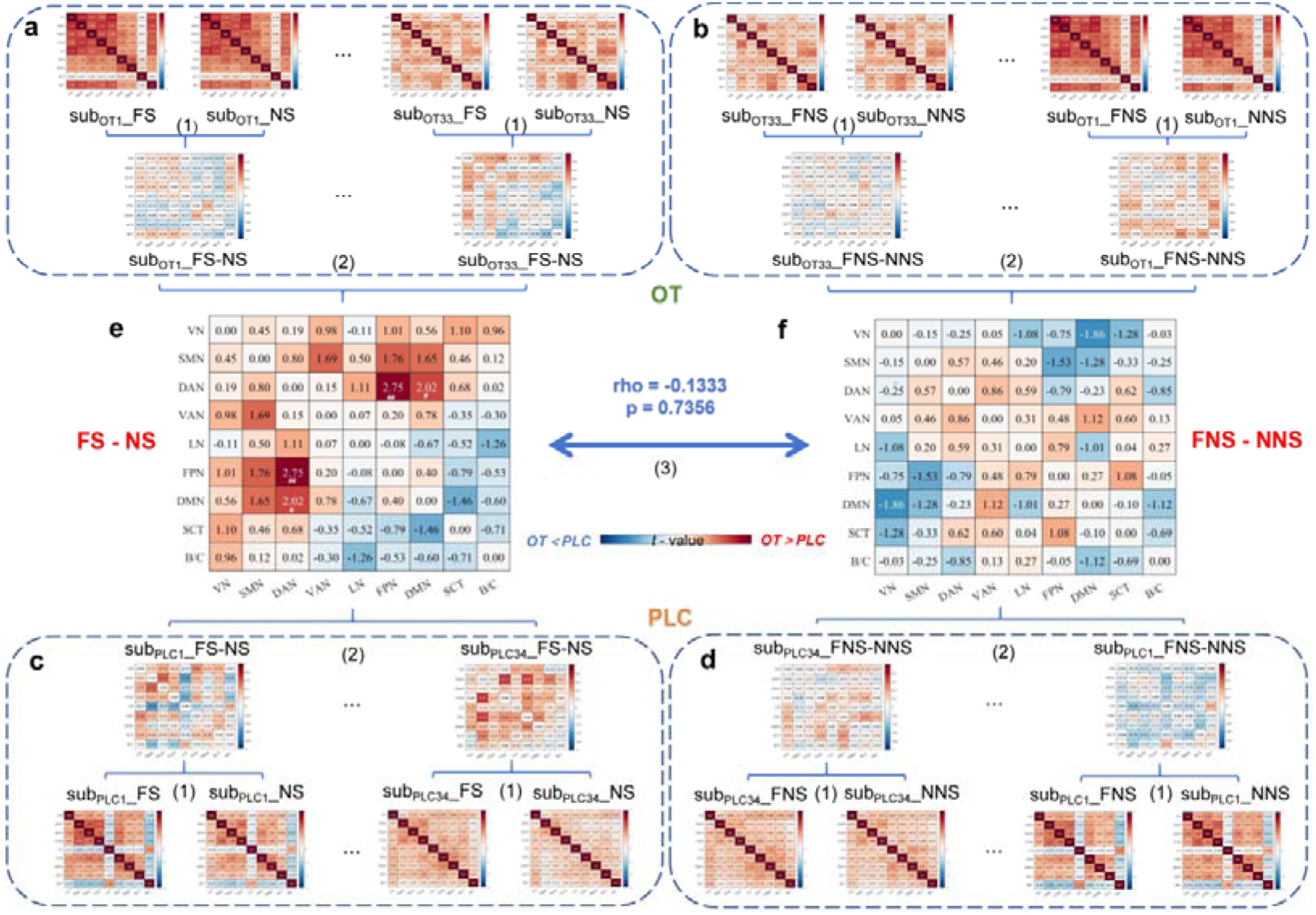
OT enhanced functional connectivity on the level of large-scale networks. **a b c d** Flow of z-value matrix calculation for the degree of response functional connectivity under specific contrasts. (1) The Fisher r-z matrices of each condition for each participant were subtracted to yield a new matrix of z-value corresponding to the contrast (e.g. z_FS_ - _NS_ = z_FS_ - z_NS_). (2) New matrices of z-value corresponding to a specific contrast were generated for each participant in each group, which were used as features in a two-sample t-test to derive the between-group differences under the specified contrast. **e** OT significantly strengthened the functional connectivity between FPN and DAN as well as DMN and DAN in the fear social-related contrast f but not in the fear non-social-related contrast. OT oxytocin, PLC placebo, FS fear social, FNS fear non-social, NS neutral social, NNS neutral non-social, VN visual network, SMN somatomotor network, DAN dorsal attention network, VAN ventral attention network, LN limbic network, FPN frontoparietal network, DMN default mode network, SCT subcortical network, B/C brainstem and cerebellum. ##pun-corrected < 0.01, # pun-corrected < 0.05.

### OT modulated whole-brain functional connectivity in the social fear context

We further conducted exploratory analyses to examine whether OT changes brain-wide communications. Results showed that while under PLC only few positive connections increased activity during the FS versus NS contrast (**Fig. 4a**), OT induced a more widespread connectivity between several brain regions (**Fig. 4b**). Under the FNS versus NNS contrast, the PLC group exhibited wide-spread connectivity (**Fig. 4c**), with no obvious changes following OT (**Fig. 4d**). Further Mantel tests and Welch’s t-test were conducted on the t-value matrices (FS - NS and FNS - NNS) between both groups showed significant modulating effects of OT under both contrasts (rho_mantel test_ = 0.0335, *t*_(1,65)_ = 9.05, *p* < 0.0001 under FS - NS, rho_mantel test_ = 0.208, *t*_(1,65)_ = -2.51, *p* = 0.012 under FNS - NNS), particularly under the fear social contexts.

**Fig. 4.**
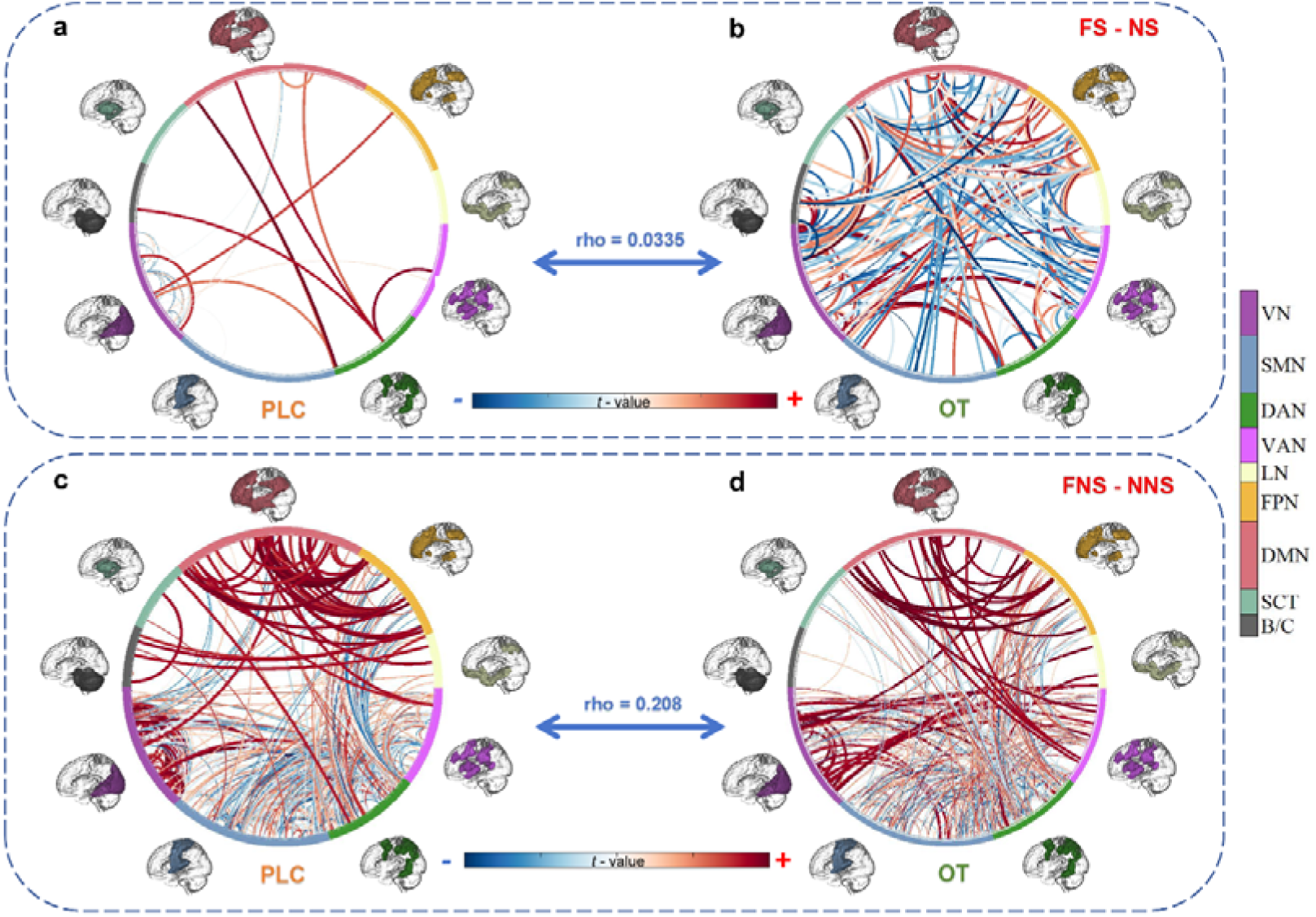
Comparison between treatment groups for whole-brain functional connectivity during social and non-social fear separately. Under fear social contexts, a PLC group showed a sparse connectivity between brain regions b the OT group exhibited stronger functional interactions at the whole-brain level. In contrast, under the fear non-social context c PLC and d OT groups showed similar whole-brain functional connectivity patterns. OT oxytocin, PLC placebo, FS fear social, FNS fear non-social, NS neutral social, NNS neutral non-social, VN visual network, SMN somatomotor network, DAN dorsal attention network, VAN ventral attention network, LN limbic network, FPN frontoparietal network, DMN default mode network, SCT subcortical network, B/C brainstem and cerebellum.

### OT attenuates the response of a synergistic brain connectivity- and activity-based signature for subjective fear (CAFE)

To comprehensively determine and independently validate the social-specific effects of OT, we utilized a recently developed neuromarker that accurately tracks subjective fear in naturalistic contexts by synergistically capitalizing on activity and connectivity^46^. The CAFE successfully determined fear versus no-fear brain responses in the entire sample (including both OT and PLC samples) with high accuracy (FS vs. NS: accuracy = 97±2.1%, area under the curve (AUC) = 1.00, P < 0.0001; FNS vs. NNS: accuracy = 100±0%, AUC = 1.00, P < 0.0001; two-alternative forced-choice test, **Fig. 5a**). This further validates the potential of the CAFE to accurately track subjective fear in dynamic contexts. In line with the univariate activation analysis and functional connectivity analysis showing that neural distinctions between treatment were only evident in the social fear contexts(FS-NS contrast), the whole-brain MVPA results revealed that the significant classification of PLC and OT treatments was demonstrated a 66% accuracy in the FS-NS contrast (mean*+*SD of accuracy = 66*±*5.8%, *p* = 0.02 , area under the curve (AUC) = 0.62, **Fig. 5b left panel**), but not in the FNS-NNS contrast (mean*±*SD of accuracy = 63*±*5.9%, *p* = 0.06, AUC = 0.60, **Fig. 5b right panel**).

**Fig. 5.**
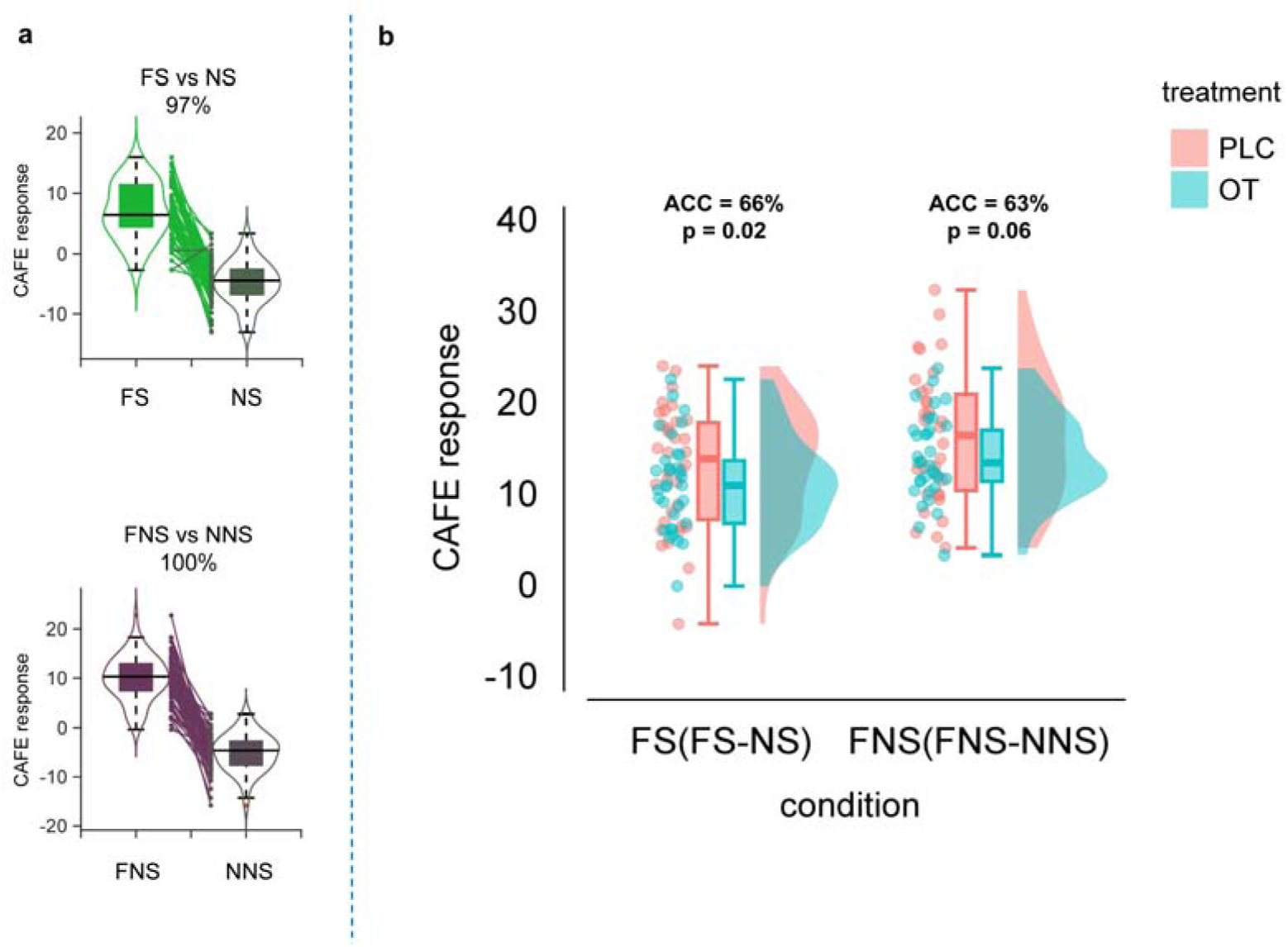
CAFE classification results. a CAFE successfully classify fear with neutral in entire group with high accuracy. b Treatment classification results of CAFE response in social and non-social fear contexts. OT oxytocin, PLC placebo, FS fear social, FNS fear non-social, NS neutral social, NNS neutral non-social.

## Discussion

The present study determined the acute behavioral and neural effects of a single intranasal OT administration on subjective fear experience in a naturalistic context that resembles dynamic fear processing in social and non-social contexts in a close-to-real life setting. On the behavioral level, OT selectively decreased the experience of subjective fear in social fear contexts, supporting a fear-regulating potential of OT under dynamic and ecologically valid social conditions. In the meanwhile, neural effects of OT were also primarily observed in social fear contexts, such that OT enhanced regional activation in the left MCC and its communication with the contralateral amygdala specifically during fear in social contexts with both neural indices exhibiting a negative association with subjective fear following OT but not PLC. Exploratory network-level analyses further revealed that OT significantly enhanced the functional connectivity between the DAN and the FPN/DMN, respectively, and brain-wide communication selectively in social fear contexts. OT additionally inhibited the CAFE response in social fear contexts but not in non-social fear contexts, which independently validates that OT regulates the whole-brain expression of fear under ecologically valid conditions and highlights the potential of multivariate neuroaffective signatures to facilitate treatment evaluation utilizing precision-pharmaco-neuroimaging^13,69^. Together, the findings indicate that a single dose of intranasal OT has the potential to decrease fear in close-to-real life settings by enhancing lMCC activation and its communication with the amygdala while concomitantly affecting network-level communication of the DAN with brain networks involved in regulatory control (FPN ^70,71^) and social processing (DMN ^72,73^) as well as brain-wide functional communication and whole-brain fear expressions.

Several previous studies in rodents and humans suggested a fear- and anxiety-reducing potential of OT, however, results remained inconsistent. Studies have described a promising fear- and anxiety-reducing neurofunctional profile of OT across species, yet the neural effects on amygdala-related circuits were often observed in the absence of changes in subjective fear experience or under experimental laboratory settings with limited ecological validity ^15,17–20,74^.

In the context of increasing attempts to enhance the ecological validity of neuroimaging research using dynamic movie stimuli ^46,47,75^, we here combined naturalistic fMRI employing fear-inducing movie clips with a pre-registered randomized double-blind placebo-controlled OT-administration trial. Across pilot experiments and treatment studies, the procedure induced a highly immersive and comparably strong experience of fear under close-to-real life conditions. Importantly, OT decreased subjective fear experience in the more naturalistic context with moderate effect size in the social context. Although the pronounced fear-reducing effects of OT in the social context may partly reflect the generally more moderate fear experience reported by our participants in this context, pronounced effects of OT in social contexts have been conceptualized in a number of overarching theories and have been reported in several previous studies 21,24,76 including studies reporting decreased subjective anxiety 77. The latter hypothesis on the social-specific effects of OT is also mirrored in the brain functional findings, such that the neurofunctional effects were selectively observed during fear experiences in social contexts.

With respect to regional activation, OT increased the activation of the left middle cingulate cortex (lMCC). The MCC represents a highly integrative hub that has been involved in both, fear evaluation ^5,78–80^ as well as the cognitive control of negative affective states including fear ^81,82^. Concomitantly, OT enhanced functional connectivity from the lMCC to the contralateral amygdala (rAmy), a pathway that has been previously involved in implementing top-down inhibitory control over excessive fear-related amygdala engagement ^78,80,83^. Effects of OT on this circuitry have been previously reported in the context of reduced subjective anxiety in individuals with PTSD 68 and in response to static fearful faces in patients with generalized social anxiety disorders ^36^. Exploring brain-behavior associations further revealed that OT established a negative association between higher activity and connectivity in this circuit and subjective fear, underscoring the behavioral relevance of the OT-induced neurofunctional changes. Together, our findings suggest that OT exerts its fear-regulating effects under naturalistic social fear contexts via enhancing MCC engagement and control over the amygdala.

On the large-scale network level, OT significantly enhanced the functional connectivity of the DAN with the FPN as well as the DMN in the social fear contexts. The DAN is centered around the intraparietal sulcus and the frontal eye fields and plays a pivotal role in top-down or goal-directed control of attention ^84–86^, while the FPN supports goal-directed actions and cognitive control ^70,87,88^ and the DMN supports self-referential ^89,90^ and social processing ^72,73^, with more recent findings additionally suggesting a role in emotional experience in dynamic naturalistic contexts ^91,92^. While studies have demonstrated the effects of OT on the interaction between these large-scale networks during the task-free state 93,94, the present findings underscore the behavioral relevance of these effects of OT in terms of regulating emotional processes in social contexts. Together with previous studies suggesting an association between dysfunctional interaction between these large-scale networks and emotional dysregulations in anxiety and fear-related disorders ^51,90^, the present findings support a potential therapeutic potential of OT for these disorders 51,95,96.

Network-level analyses that explored the effects of OT on the fine-grained level of brain-wide connectivity patterns suggested that OT may broadly facilitate brain-wide exchange of information within and across networks particularly between the FPN and DMN in social fear contexts. The results align with recent findings suggesting that emotional processes such as fear and anxiety are processed in distributed networks and their interactions 46,53,97 as well as the distributed network-level properties and effects of the OT signaling system ^60,66,98^. Findings on the whole-brain effects were additionally validated using a precision pharmaco-neuroimaging approach which capitalizes on MVPA-based decoding. To this end we employed an independently developed fMRI-based marker that accurately tracks subjective fear in naturalistic contexts (CAFE^46^). The marker precisely tracked subjective fear under ecologically valid contexts in both groups, yet showed an attenuated reactivity in the OT group, in particular in the social fear context. Independent whole-brain-level MVPA analysis indicated that OT has the potential to diminish distributed fear pattern. These findings underscore that OT may exert its effects on human emotion processing under complex naturalistic conditions via regulating extensive brain wide interactions. Together with current reconceptualizations proposing that emotional processes are represented in brain-wide distributions^97,99^ and that mental disorders are not related to dysfunctions of a single brain region or pathway but rather dysregulated brain-wide communication ^51,90,100^, these findings further underscore the therapeutic potential of the OT signaling system.

Findings have to be considered in the context of limitations of the present design, including (1) to avoid variations in the effects of OT related to sex ^101,102^, we only focused on male participants and future studies need to test generalization to females, (2) the study employed a proof-of-concept design in healthy subjects and future studies need to validate the effects of OT in populations with pathological fear and anxiety, (3) despite extensive pre-study testing the videos of the social and non-social category differed in terms of the level of fear induced which partly limits conclusions regarding social-specific effects of OT, (4) the videos included homogenous brief episodes of fear and future studies need to examine effects of OT on fear during longer time scales and longer episodes with dynamic fear ^46,47^.

Taken together, this study demonstrated that OT can reduce subjective fear experience under naturalistic social fear conditions via enhancing lMCC engagement and its regulatory control over the amygdala as well as enhancing communication between large-scale networks and brain-wide communication. These findings consistently indicate a high translational therapeutic potential of OT to regulate fear in dynamic and close-to-real life environments. Effects were pronounced or restricted to the social contexts, supporting the role that OT may play in social contexts, and rendering OT as a promising treatment for social context-specific fear experiences, e.g. in social anxiety disorders. Findings indicate that OT may be used as a novel therapeutic strategy to regulate excessive subjective fear symptoms in mental disorders ^7,17,35^ or may help to prevent the development of these disorders during early stages or promote long-term recovery ^94^ via improving the quality of life, and reduce the social and economic burden related to fear-related disorders.

## Methods

### Participants of the fMRI study

Sixty-nine right-handed male participants with normal or corrected-normal vision were recruited from the University of Electronic Science and Technology of China. The sample size was determined using G*Power v.3.1 103,104 indicating that 62 participants were required to detect a significant effect on the behavioral level (*f* = 0.25, *α* = 0.05, *β* = 0.80, 2 *X* 2 *X* 2 mixed ANOVA interaction effect, with treatment as between-subject factor, and emotion and social as within-subject factors). The present study focused on male subjects to reduce variance related to sex differences in the effects of OT ^101,102^ and adhered to validated exclusion criteria (details see Supplementary Methods). The final sample size was n = 67 (mean *±* SD, age = 21.03 *+* 1.95 years) due to two data from two participants being excluded (withdraw due to personal reasons, technical MRI issues). All participants provided written informed consent, protocols and the primary outcomes (fear ratings and brain activity) were pre-registered on Clinical Trials.gov (clinicaltrials.gov/ NCT05892939), approved by the ethics committee at the University of Electronic Science and Technology of China (UESTC-1061423041725893) and in line with the latest Declaration of Helsinki.

Using a double-blind randomized, placebo-controlled, between-subject pharmacological fMRI design, participants were randomly assigned to receive either a single intranasal dose of oxytocin (OT, 24IU-Sichuan Defeng Pharmaceutical Co. Ltd, China) or a placebo (PLC, same ingredients i.e. sodium chloride and glycerin but without the peptide). The spray bottles were identical and dispensed by an independent researcher based on a computer-generated randomization sequence to ensure double-blinding. The investigators involved in data acquisition and analyses were blinded for group allocation. Following recent recommendations ^105–107^, the fMRI assessment began 45 minutes after treatment administration.

To control for pre-treatment between-group differences, participants completed a series of mood and mental health questionnaires (as detailed in **Table 1**). Participants were asked to guess which treatment they had received after the experiment (with a non-significant treatment guess *χ*^2^ = 0.39, *p* = 0.53, confirming successful double-blinding). Additionally, participants were required to complete a surprise recognition task after the experiment that involved presenting stills of the video clips shown during fMRI and new clips to test the attentive processing of the stimuli. All participants achieved a re-recognition accuracy of over 75% and no significant difference in performance was observed between the two treatment groups (*t*_(1,65)_ = -0.693, *p* = 0.491).

### Naturalistic fear induction paradigm

The task began 45 minutes after the treatment intervention (see **Fig. 6a**). During the naturalistic fear induction paradigm, participants were instructed to watch a series of video clips attentively and rate their level of fear experience after each clip. The clips lasted 25 seconds and were followed by a 1-3 second jittered fixation cross to separate the stimuli from the rating period. During the 5-second period following each stimulus, participants were asked to report their level of subjective fear on a 9-point Likert scale, with 1 indicating no fear and 9 indicating very strong fear. This was followed by a jittered inter-trial interval of 9-11 seconds, during which a fixation cross was presented (see **Fig. 6b**). A total of 32 short video clips were presented over 2 runs, with each run consisting of 16 randomly presented clips, half of which were neutral and half of which were fear-inducing clips.

**Fig. 6.**
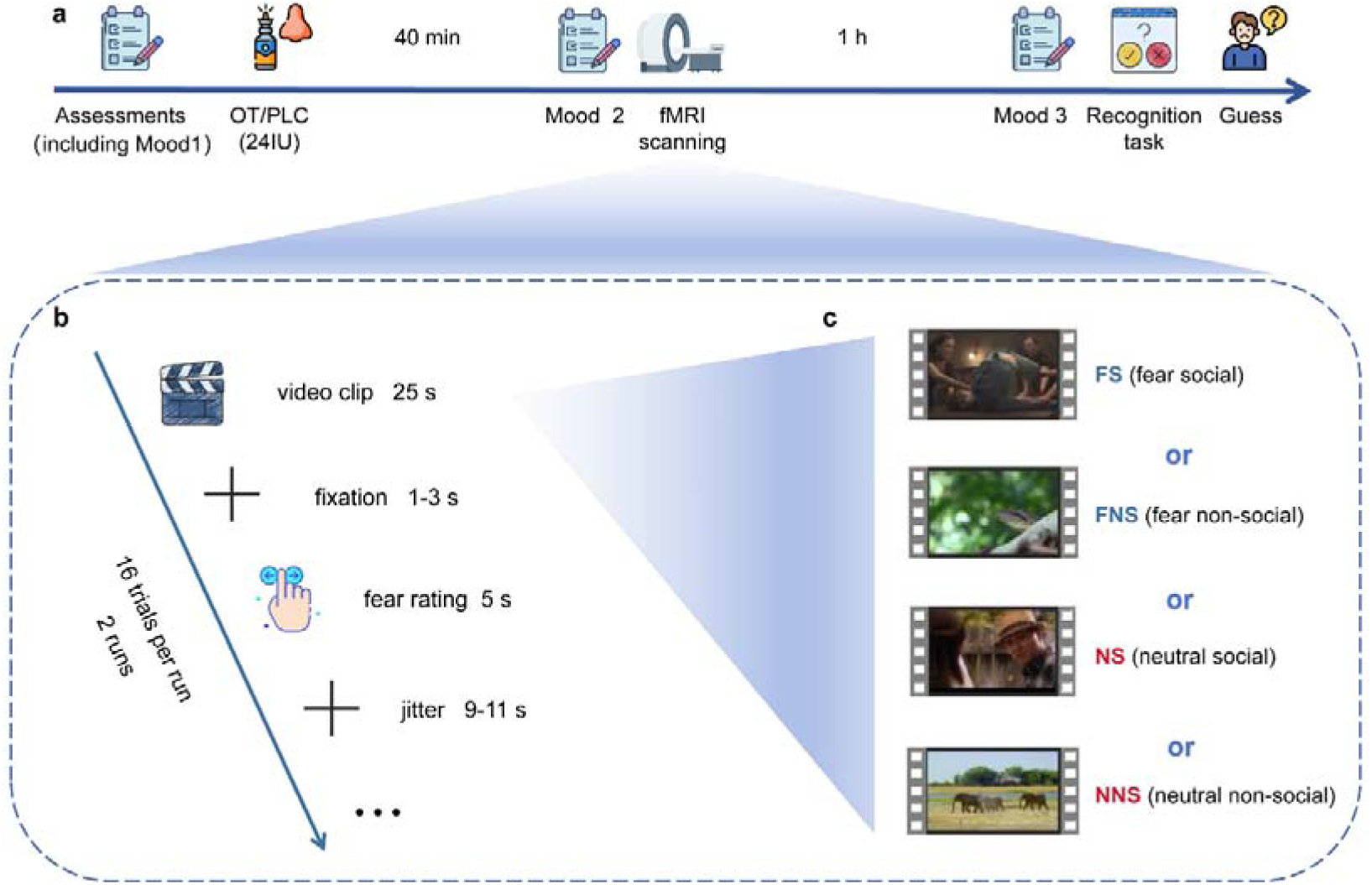
Experimental timeline, paradigm and schematic depiction of stimuli. **a** The timeline of the whole experiment. **b** Naturalistic fear induction and rating paradigm. **c** Example stimuli. In each run, 4 types of video clips were presented (FS, FNS, NS, NNS) according to a randomized order. Abbreviations: OT oxytocin, PLC placebo, fMRI functional magnetic resonance imaging, FS fear social, FNS fear non-social, NS neutral social, NNS neutral non-social. License: Images in 1a and 1b were obtained from Flaticon.com under the free license with attribution.

Given ongoing discussions about the social-specific effects of OT ^108,109^, we carefully pre-selected a balanced set of stimuli that included social as well as non-social fearful or neutral stimuli, respectively. This led to a set of four stimulus categories (fear social clips-FS, fear non-social clips-FNS, neutral social clips-NS, neutral non-social clips-NNS, see **Fig. 6c**). Before the OT experiment, the video clips were carefully evaluated and selected based on the results of a behavioral pilot study in an independent sample (pilot study, n = 20; 10 females; age = 22.65 ± 2.01). For detailed characterization of the video clips and details of the pilot study, see Supplementary Methods. Based on the results from the pilot study the video clips used for the naturalistic fear induction fMRI paradigm were selected from a larger database. Key selection criteria for the video clips were the ability to induce strong, emotion-specific and consistent feelings of subjective fear or a neutral emotional state, respectively (see Supplementary Methods). Based on the results of the corresponding ratings in the pilot study (see **Fig. S2**), 8 complimentary video clips were selected for each of the four categories. The task was programmed in Python 3.7 using the PsychoPy package (version 2022.2.4).

### Effects of OT on subjective fear experience

To determine the effects of OT on subjective fear in naturalistic contexts and whether they vary as a function of social context, we computed a repeated measures ANOVA with the between-subject factor treatment (OT vs PLC) and the within-subject factors emotion (fear vs neutral) and social (social vs non-social) and the dependent variable fear ratings during fMRI. Analyses were conducted using GraphPad Prism 9.0 (GraphPad Software, Inc., La Jolla, CA), and corresponding effect sizes were computed using JASP 0.18.3.0 (JASP Team, 2019; jasp-stats.org). All ANOVAs and t-tests used false discovery rate (FDR) ^110^ recommended by Prism for multiple comparison correction.

### MRI data acquisition and preprocessing

MRI data were collected on a 3.0-T GE Discovery MR750 system (General Electric Medical System, Milwaukee, WI, USA) and preprocessed using fMRIPrep 21.0.0111 (RRID: SCR_016216), a Nipype 1.6.1 based tool that integrates preprocessing routines from different software packages and SPM 12 (Statistical Parametric Mapping; http://www.fil.ion.ucl.ac.uk/spm/; Wellcome Trust Centre for Neuroimaging, detailed in Supplementary Methods).

### Effects of OT on fear-related brain activity

To examine the effects of OT on fear-related neural activity in naturalistic contexts, a GLM-based model was established in SPM 12. The model incorporated five regressors: FS, FNS, NS, NNS, and the rating period. We included the 24 head motion parameters as covariates of no interest.In line with previous research47, we removed variance associated with the mean, linear and quadratic trends, the average signal within anatomically-derived CSF mask, the effects of motion estimated during the head-motion correction using an expanded set of 24 motion parameters (six realignment parameters, their squares, their derivatives, and their squared derivatives) and motion spikes (FMRIPREP default: FD *>* 0.5mm or standardized DVARS *>* 1.5)^112^. Key first-level contrasts of interest were modeled on fear in social and non-social contexts separately (i.e., [FS - NS] or [FNS - NNS]) as well as their interaction (i.e., [FS - NS] - [FNS - NNS]). In line with the behavioral analyses, the effects of OT were determined using an ANOVA with the factors of emotion and social contexts. Analyses were implemented within a partitioned error ANOVA approach that subjected the first level interaction contrast to a voxel-wise two-sample t-test with the factor treatment group modelled on the second level (i.e., [OT - PLC] or [PLC - OT]). Based on our hypothesis and previous studies reporting that fear is strongly associated with the engagement of amygdala, cingulate cortex (particularly middle cingulate cortex, MCC), insula, and ventromedial prefrontal cortex (vmPFC) and that oxytocin mediates its anxiolytic effects via these regions 14,46,76,78,113,114. Our analyses focused on atlas-derived bilateral masks for the amygdala (a combined mask generated from the Melbourne subcortex atlas^115^ and the extended amygdala mask from the reinforcement learning atlas^116^), MCC (combined bilateral FPN_Cing_1 and FPN_Cing_2 from the Schaefer atlas, 400 ROI version^67^), insula (bilateral VAN_FrOperIns from the Schaefer atlas) and vmPFC (combined bilateral FPN_PFCv and DMN_PFCv from the Schaefer atlas). The masks were utilized in separate small volume corrected (SVC) and family-wise error (FWE) peak level corrected ^117^ analyses.

To further disentangle complex interaction effects involving treatment, parameter estimates were extracted using an atlas-based structural mask from the Schaefer atlas representing the left MCC^67^. Estimates were extracted using Marsbar (http://marsbar.Sourceforg-e.net/). The group differences based on the extracted BOLD signals in each of the separate conditions were conducted using two-sample t-test and presented for display purpose.

Additionally, to determine associations between the behavioral (subjective fear) and neural effects of OT (both activation and functional connectivity between specific brain regions, parameter estimates were derived using independently defined structural masks (left MCC for activation and right amygdala for functional connectivity from the Schaefer atlas and Melbourne subcortex atlas^115^ and the extended amygdala mask from the reinforcement learning atlas^116^)). We conducted Pearson correlation analysis using using GraphPad Prism 9.0 (GraphPad Software, Inc., La Jolla, CA), It should be noted that the correlation analyses were conducted for exploratory purpose that need to be interpreted with caution.

### Effects of OT on functional communication of the regions identified

We further examined whether the regions showing altered regional activation following OT also changed their network-level communication via a general Psychophysiological interaction (gPPI) analysis. gPPI examines functional connectivity between brain regions that is contingent on a psychological context. We used the Generalized PPI toolbox ^118^ as implemented in SPM 12, to conduct gPPI. This method includes additional task regressors to reduce the likelihood that the functional connectivity estimates are driven simply by co-activation and has been widely used in similar studies ^119,120^. Based on our results, OT enhanced activation in the left middle cingulate cortex (lMCC) under the three-way interaction (treatment *X* emotion *X* social), we therefore computed the functional communication of this region (using an 8mm radius which centered in peak Montreal Neurological Institute (MNI), x, y, z = -7, -3, 30) with other voxels in the whole brain under the two-way interaction of emotion and social, and a two-sample t-test was conducted to confirm the treatment effect of OT. Based on previous studies reporting effects of OT on the functional communication of the amygdala ^101,102^, separate atlas-derived bilateral masks^67^ for the amygdala, MCC, insula and vmPFC were used for SVC analyses with FWE correction 117 applied at peak level.

Marsbar was used to extract the lMCC - rAMY functional coupling using independent rAMY mask^67^ to further disentangle complex interaction effects involving treatment. The extracted functional connectivity signals were then subjected to compare the treatment effect under four conditions by using two-sample t-tests separately.

### Effects of OT on large-scale brain network functional connectivity

Given that several studies reported the effects of OT on large-scale network organization of the brain ^93,121^, we examined OT effects on the whole-brain network level. To this end, time-series of each region of interest (ROI) was extracted according to the template including 463 ROIs across the whole brain (see Supplementary Methods), resulting in BOLD time series of the size of *r X t* (*r* was the number of ROIs and *t* was the BOLD time series length) for each subject per run. A matrix of size *r X r* was obtained by calculating Pearson’s correlation between the average BOLD time series of each pair of ROIs. Additionally, Fisher’s z-transform was applied to improve the normality of the correlation coefficients. The brain regions can be divided into nine networks (visual network, VN; somatomotor network, SMN; dorsal attention network, DAN; ventral attention network, VAN; limbic network, LN; frontoparietal network, FPN; default network, DMN from Yeo 7 networks^122^; subcortex network, SCT115; brainstem and cerebellum network, B/C56) based on their functional divisions, and the neural activities of each network’s regions were averaged to calculate network functional connectivity.

To extract neural activity time series for a specific condition, we integrated the BOLD time series of the different runs into one time series and modelled the onset time and duration of the stimuli per condition. This resulted in four *r X d* matrices (where *r* is the number of ROIs and *d* is the BOLD time series length of each condition). Given the typical hemodynamic response function (HRF) lag of 6 seconds46, we shifted the onset times by 3 TRs – corresponding to 6 seconds - of the BOLD time-series data relative to the clip stimuli onsets. Additionally, we obtained the *r X r* matrix between ROIs and the *n X n* matrix (where *n* is the number of networks) between brain networks for different conditions using Pearson correlation analysis.

After performing Fisher-Z transformation on the network functional connectivity matrices for different conditions, matrices of Z-values corresponding to specific conditions (Z_FS_\Z_FNS_\Z_NS_\Z_FNS_) are obtained. It is important to note that r values are not additive or subtractive, but Z scores are summative. Matrix of Z-values corresponding to particular contrasts (e.g. Z_FS-NS_) can be computed by subtract the matrices of Z-values corresponding to different conditions (e.g. Z -Z)^123,124^. Therefore, each participant in both groups would have their own subtracted Z-values matrix under specific contrasts (Z_FS-NS_\Z_FNS-NNS_). We then conducted two-sample t-tests on each pair of network-level functional connectivity to determine the treatment effect.

### Effects of OT on whole-brain functional connectivity

To further investigate the impact of OT on a finer spatial scale, we analyzed whole-brain functional connectivity (used whole-brain ROIs functional connectivity matrices for further computation) using paired t-tests for key contrasts (FS - NS and FNS - NNS) separately for the two groups. We applied FDR correction (*p_FDR-corrected_* < 0.05) to identify the most relevant functional connections for each contrast. A Mantel test ^125,126^ was performed on the FDR-corrected functional connectivity matrices of the two groups, under specific contrasts, to assess their similarity. Furthermore, to investigate treatment-specific effects on functional connectivity strength at the whole-brain level, we transformed the upper triangular data of the *t*-value matrices of the two groups under specific contrast into two (*r X* (*r -* 1)) / 2 by 1 feature vectors, and performed Welch’s t-test between them.

### Effects of OT on synergistic brain connectivity- and activity-based signature for subjective fear (CAFE)

We applied a recently developed whole-brain multivariate neuromarker for fear in naturalistic contexts (CAFE46) which synergistically capitalizes on brain connectivity- and activity-based signature to accurately track subjective fear in dynamic naturalistic contexts^46^ to independently validate effects of OT on the whole-brain expression of fear (for a brief description of the development of the CAFE please see also Supplementary Methods). First, we calculated the mean beta value vector(one for each region; n=463 in total, see Supplemental Methods in detail) and functional connectivity matrix (463 *X* 463) for each subject under the four conditions (FS\FNS\NS\NNS) and applied dot-product the integrated one-dimensional vector (n = 107416 features in total) with the CAFE weights to calculate the CAFE response for each subject under each of the four conditions. Second, in line with the other analyses we build the contrasts of interest (FS-NS; FNS-NNS) and calculated CAFE response of each contrast. To quantify the classification performance of the model (classification performance between PLC and OT under same contrast condition), Receiver Operating Characteristic (ROC) analysis was conducted.

## Data Availability

All data produced in the present study are available upon reasonable request to the corresponding author

## Acknowledgement

This work was supported by the China MOST2030 Brain Project (Grant No. 2022ZD0208500-BB), the National Natural Science Foundation of China (Grant No. 82271583-BB), the National Key Research and Development Program of China (Grant No. 2018YFA0701400-BB), the Natural Science Foundation of Sichuan Province (Grant number 2022NSFSC1375-WZ) and a start-up grant from The University of Hong Kong. Disclaimer: Any opinions, findings, conclusions or recommendations expressed in this publication do not reflect the views of the Government of the Hong Kong Special Administrative Region or the Innovation and Technology Commission.

## Author contributions

KF and BB1 designed the study. KF, ZZ and DL conducted the experiment and collected the data. KF, SX, WZ, ZL performed the data analysis with the help of TX, YZ, XZ, CL, JW, LW, JH and BB^2^. KF and BB^1^ wrote the manuscript. SX, FZ, KK, TX, YZ, DY, ZL and WZ critically revised the manuscript draft.

Note: BB^1^, Benjamin Becker; BB^2^, Bharat Biswal.

