## Supplementary material for "Oxytocin reduces subjective fear in naturalistic social contexts via enhancing top-down middle cingulate amygdala regulation and brain-wide fear representations": SI for OT-fear paper (from another version of this preprint: version 2, 2025-03-04)

**Affiliations**

### Supplemental Methods

#### Participants

Exclusion criteria of participants comprised a history of head injury, color blindness, current or regular substance or medication use, current or history of medical or psychiatric disorders, and any contraindications for oxytocin or MRI.

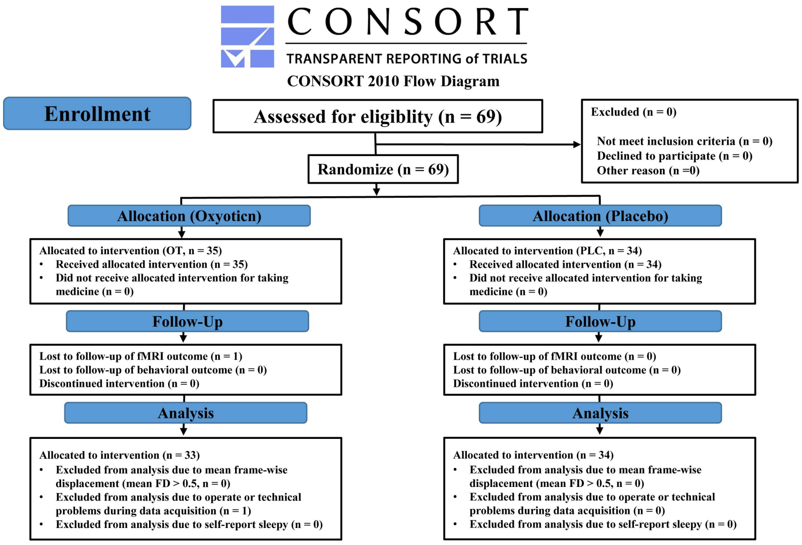


**Fig. S1 CONSORT Flow Diagram.**

#### MRI acquisition and preprocessing

Functional MRI data was acquired using a T2*-weighted echo-planar imaging (EPI) pulse sequence (repetition time = 2000 ms, echo time = 30 ms, 36 slices, slice thickness = 3.8 mm, no gap, field of view = 200 × 200 mm, resolution = 64 × 64, flip angle = 90°, 3.125 × 3.125 × 3.8 mm voxels, axial scan plane). To improve spatial normalization and exclude participants with apparent brain pathologies a high-resolution T1-weighted image was acquired using a 3D spoiled gradient recalled (SPGR) sequence (176 slices, repetition time = 8.22 ms, echo time = 3.15 ms, field of view = 256 × 256 mm, resolution = 256 × 256, flip angle = 8°, 1 × 1 × 1 mm voxels).

The MRI images preprocessing included 1) Spatial normalization to the ICBM 152 Nonlinear Asymmetrical template version 2009c was performed through nonlinear registration with the antsRegistration tool of ANTs v2.3.3 [1], using brain-extracted versions of both T1w volume and template. 2) Brain tissue segmentation of cerebrospinal fluid (CSF), white matter (WM) and gray matter (GM) was performed on the brain-extracted T1w using FAST (FSL v6.0.5.1) [2]. 3) Before the automated preprocessing, 5 initial volumes of fMRI data were removed to allow for image intensity stabilization. 4) Next, functional data was slice time corrected using 3dTshift from AFNI [3] and motion corrected using mcflirt (FSL v6.0.5.1). This was followed by 5) co-registration to the corresponding T1w using boundary-based registration [4] with six degrees of freedom, using FLIRT (FSL). 6) Motion correcting transformations, 7) BOLD-to-T1w transformation and 8) T1w-to-template (MNI) warp (interpolated to 2 mm isotropic voxels) were concatenated and applied in a single step using ants ApplyTransforms (ANTs v2.3.3) using Lanczos interpolation.

Preprocessed images were spatially smoothed using an 8-mm full-width at half maximum

(FWHM) Gaussian kernel in SPM 12.

#### Definiton of ROIs

The template utilized in this study comprises 400 cortical regions from the Schaefer atlas [5], 34 subcortical regions from the Melbourne subcortex atlas [6], and the reinforcement learning atlas (extended amygdala and hypothalamus) [7] and periaqueductal gray (PAG), brainstem, and cerebellar regions (n = 29) [8], resulting in a total of 463 regions.

#### Video clips materials evaluation procedure

The participants were required to watch 40 short video clips attentively and rate their feelings using a series of 9-point Likert scales while watching the videos on five dimensions after watching each video clip (see **Fig. S2**). The five rating scales included arousal (where 1 represents very calm; 9 indicatesextremely arousing), valence (where 1 represents very positive and pleasurable; 9 represents very negative), fear (where 1 represents no fear at all; 9 represents extreme fear), social (where 1 represents exceptionally low levels of sociability, with no social context at all; 9 indicates exceptionally high levels of sociability, with high levels of social context and interactions), and consistency (where 1 extermely low emotional consistency of the video clip; 9 represents a high level of emotional consistency across the movie clip).

To screen the stimulus materials that matched our experimental design, we sorted all the videos on each of the five rating dimensions from highest to lowest and selected the video clips for the subsequent MRI experiments (8 clips for each of the 4 conditions FS\FNS\NS\NNS). To further verify that the selected video clips could effectively evoke the corresponding emotions, we performed a series of repeated measures ANOVAs on each of the five rating dimensions of the selected stimulus materials, respectively.

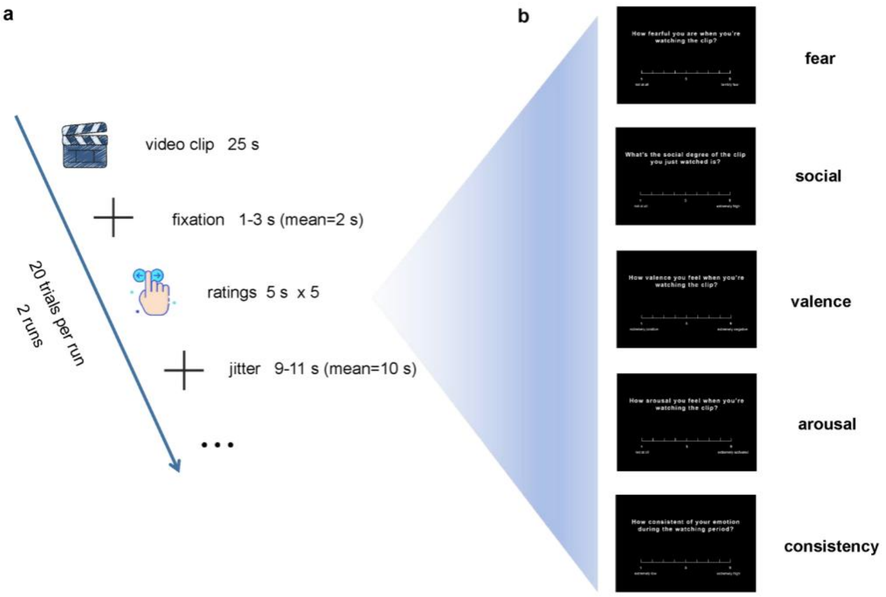


**Fig. S2 Video clips materials evaluation procedure.**

#### Key exclusion criteria for clip materials selection

(1) A lack of fear-specificity as operationalized by more than 1 participant (5%) reporting induction of non-fear negative (e.g., disgust and sad) or positive (e.g., happiness and joy) emotions during the movie, and (2) low emotional consistency over the duration of the clip,

operationalized as a mean consistency rating ≤ 6.5 on a 9-point Likert scale with 1 indicating that the intensity of the elicited emotion was very inconsistent throughout the video clip and 9 indicating that the intensity of the elicited emotion was very consistent throughout the video clip.

#### Supplemental Results

##### The selected video clips were sufficient to induce corresponding emotions on both behavioral and neural level.

Compared to neutral clips, fear video clips (FS and FNS) yielded considerably higher fear ratings, and the main effects of valence and social was significant ($F_{(1,18)}$ = 290.6, $p$ < 0.0001; $F_{(1,18)}$ = 25.92, $p$=0.001, **Fig. S3a**), the interaction between valence and social was also significant($F_{(3,18)}$ = 42.81, $p$ = 0.0001, **Fig. S3a**). To further eliminate the possible confounding effects of valence and arousal, no main effect of social was found for either of these rating indices (all $p$s > 0.05, **Fig. S3c** and **Fig. S3e**). Social video clips (FS and NS) yeileded much higher social ratings compared to non-social ones, the main effect of social was significant ($F_{(1,18)}$ = 611.2, $p$ < 0.0001, **Fig. S3b**). In addition, all selected video clips with pretty high consistency rating scores (see **Fig. S3d**).

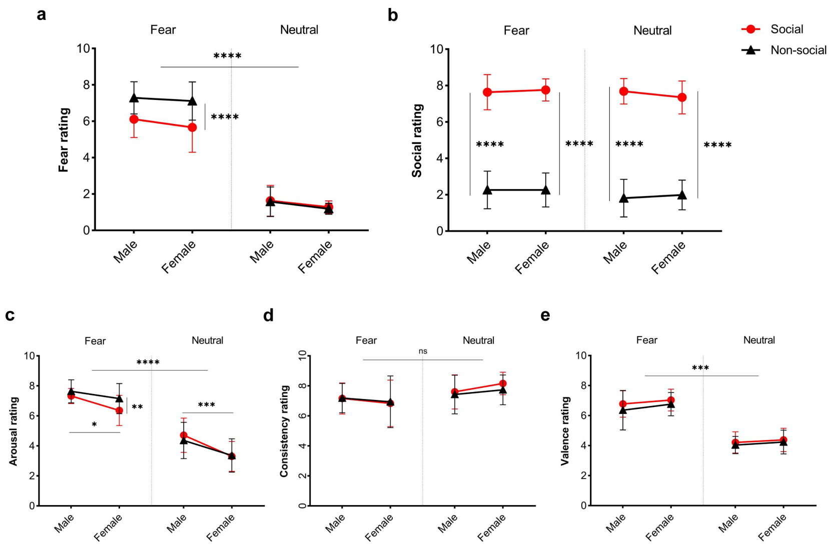


**Fig. S3 The selected stimuli for fMRI experiment were sufficient to induce corresponding emotions. a** The participants tended to rate subjective fear lower on fear social clips compared to their non-social peers in both gender, but this difference didn't survive on **e** valence rating and **c** arousal rating (only in female, so we recruited only males in the latter fMRI study to remove the possible confounding roles and gender differences). **b** The selected social stimuli all have much higher rating scores on social than non-social controls. In addition, all selected stimuli have **d** pretty high consistency rating scores.

At the neural level, fear video clips (FS and FNS) elicited greater activation in fear processing regions compared to neutral clips. In addition, the activated neural patterns in social and non-social fear contexts showed widely distributed common regions as well as some distinct areas (see **Fig. S4**). All the above results consistently suggested that the video clips we selected as the formal stimuli materials were suitable and adequate to induce corresponding emotions.

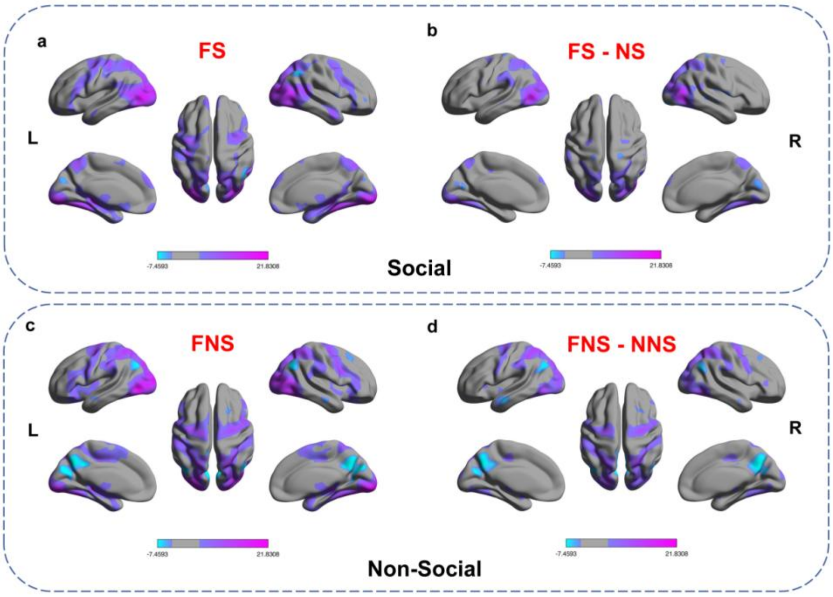


Fig. S4 Shared and distinct activated neural patterns induced by the selected stimuli for fMRI experiment under social and non-social fear contexts. All the results presented here came from only PLC group to see the overall activation induced by video clips and at peak-level unthreshold $p < 0.001$, FS fear social, FNS fear non-social, NS neutral social, NNS neutral non-social.

Brief descriptions of each video clip used in the fMRI study were also provided in **Table. S1**.

**Table. S1 Detailed information about the selected stimuli**

| Stimuli type | Number | Brief description |
|---|---|---|
| FS | FS01 | Demonstrators clash with police in standoff, scene of flames |
| | FS02 | Smoke billows from clashes between police and demonstrators |
| | FS03 | Heavy clashes as many armed men ganged up on one man |
| | FS04 | Two social gangs were fighting each other with weapons, and the scene was intense |
| | FS05 | Very strong earthquake scene, people running for their lives |
| | FS06 | Rescue after the earthquake, rescuers are passing through fast and dangerous rivers |
| | FS07 | Firefighters work to extinguish a house fire as it rages on |
| | FS08 | Firefighters work to extinguish a factory fire as it rages on |
| FNS | FNS01 | A man performing a dangerous maneuver at high altitude |
| | FNS02 | A man running fast at high altitude from a first viewpoint |
| | FNS03 | A python is strangling its prey and opening its bloody mouth to try and swallow it |
| | FNS04 | A giant python is spitting on the riverbank and crawling closer |
| | FNS05 | A giant spider crawls and feeds fast |
| | FNS06 | A close-up of a giant spider and its feeding scene |
| | FNS07 | First view of a fast ride in the very rugged outdoors |
| | FNS08 | First view of a fast ride in the very rugged outdoors with body flips |
| NS | NS01 | Two students go up and explain while the teacher and classmates listen from below |
| | NS02 | The teacher is giving a lecture and the students are listening carefully |
| | NS03 | Two schoolgirls met and sat down to have a short chat |
| | NS04 | A man and a woman were sitting on benches near the river and chatting face to face |
| | NS05 | A chemist explaining science-related work to the public |
| | NS06 | A physicist explaining science-related work to the public |
| | NS07 | Hanging out at the mall in the first place |

| | | |
|---|---|---|
| | NS08 | Hanging out at the supermarket in the first place |
| NNS | NNS01 | A herd of cows is grazing and eating milk on the grassland |
| | NNS02 | A herd of cows is grazing and resting in the pasture |
| | NNS03 | A flock of seagulls circled along the highway by the sea |
| | NNS04 | A flock of seagulls circled along the sandy beach by the sea |
| | NNS05 | Calm lake and relaxed animals |
| | NNS06 | Calm lakes and quiet woods |
| | NNS07 | Greenery swaying in the breeze and beautiful sunsets |
| | NNS08 | Pieces of wheat swaying in the breeze |
